# Influence of frailty on cardiovascular events and mortality in patients with Chronic Obstructive Pulmonary Disease (COPD): Study Protocol for a multicentre European observational study

**DOI:** 10.1101/2024.03.10.24304053

**Authors:** A Verduri, E Clini, B Carter, J Hewitt

## Abstract

**Background:** Frailty is a clinical state that increases susceptibility to minor stressor events. The risk of frailty is higher in chronic conditions, such as Chronic Obstructive Pulmonary Disease (COPD). Recent studies on COPD have shown that patients living with frailty have an increased risk of mortality. The presence of cardiovascular diseases or conditions are common in COPD and may increase the risk of death.

**Methods:** This protocol describes a European prospective cohort study of community-based people, in a stable condition with diagnosis of COPD (as defined by GOLD guidelines) across hospitals in Italy and UK. Frailty prevalence will be assessed using the Clinical Frailty Scale. At 1- and 2-year follow up, primary outcome will be the impact of frailty on the number of cardiovascular events; secondary outcomes: the influence of frailty on cardiovascular mortality, all-cause mortality, and deaths due to COPD. For the primary outcome a zero-inflated Poisson regression will compare the number of cardiovascular events at 1 year. Secondary outcomes will be analysed using the time to mortality.

**Discussion:** This multicentre study will assess the association between frailty and cardiovascular events and mortality in population with COPD. Data collection is prospective and includes routine clinical data. This research will have important implications for the management of patients with COPD to improve their quality of care, and potentially prognosis.

**Trial registration number:** NCT05922202 (www.clinicaltrials.gov).

## INTRODUCTION

Frailty is a clinical syndrome that increases vulnerability to stressors due to decline of multiple physiological reserves. People living with frailty area at higher risk of falls, disability, long hospitalisations, access to care homes, and death. [1] There are two principal approaches of identification of frailty: the deficit accumulation model and the phenotype model. [1,2] Based on these models, different tools have been created and validated to assess frailty in clinical practice. [3]

Chronic diseases such as Chronic Obstructive Pulmonary Disease (COPD) increase the risk of frailty. [3,4] Literature on frailty in COPD have been emerging in the recent years. While frailty is not synonym for ageing, it is well known that people over 65 years are more likely to live with frailty. [4] In addition, prevalence of COPD increases with age. [5] A double probability of being frail has been demonstrated in patients with COPD compared to patients without COPD in the paper of Marengoni et al. The study reported a prevalence of frailty in COPD between 9% and 64% based on the phenotype model and from 9% to 28% using different frailty assessment instruments. [6] Studies have shown that frailty can increase the risk of mortality in patients with confirmed diagnosis of COPD according to GOLD guidelines (www.goldcopd.org). [7-12]

Cardiovascular diseases share common risk factors with COPD such as ageing, active smoking, and sedentary lifestyle. COPD and CVD frequently co-exist and are associated with worse outcomes than either condition alone. Additionally, the risk of cardiovascular events is higher after COPD exacerbation. [13,14] Cardiovascular comorbidities are very common in patients with COPD and can increase the risk of death in this population. [5,15,16]

The present protocol describes a research study project on patients with COPD to investigate the influence that frailty can have on the risk of cardiovascular events in these patients. The project will assess the association between frailty and cardiovascular mortality, and the relationship between frailty and all-cause mortality and COPD-related mortality. Our team have widely collected data on frailty previously using routinely collected service evaluation level data.

The study has the goal to provide relevant data for clinical practice and management of patients with COPD to improve quality of care, and potentially prognosis, considering a more accurate characterisation of the patient living with COPD.

### Aims

To assess whether frailty correlates with cardiovascular outcomes in patients diagnosed with COPD according to guidelines. The following research questions will be addressed:

1. How many cardiovascular events (including hospital admissions due to cardiovascular cause) at 12 and 24 months in COPD patients living with frailty *vs* non-frail COPD patients. Coronary artery disease (CAD), aortic disease, peripheral artery disease (PAD), cerebrovascular disease, deep vein thrombosis (DVT) and pulmonary embolism (PE), and arrhythmias were selected as cardiovascular events. The list of ICD-10 codes related to cardiovascular events of special interest for the study protocol is shown in **Table 1**.
2. How many deaths due to cardiovascular cause at 12 and 24 months in COPD patients living with frailty vs non-frail COPD patients.
3. How many deaths due to all-cause mortality at 12 and 24 months in COPD patients living with frailty *vs* non-frail COPD patients.
4. How many deaths due to COPD at 12 and 24 months in COPD patients living with frailty *vs* non-frail COPD patients.
5. Prevalence of frailty in COPD.

**Table 1.**

| **Table 1** | **ICD-10 code** |
| --- | --- |
| **Coronary artery disease (CAD)** |  |
| Angina | **I20** |
| Acute Myocardial Infarction (AMI) | **I21** |
| Heart failure | **I50** |
| **Aortic disease** |  |
| Aortic aneurysm | **I71.2** |
| Abdominal aneurysm | **I71.4** |
| **Peripheral artery disease (PAD)** | **I73** |
| **Cerebrovascular disease** |  |
| Ischemic stroke | **I63** |
| Intracranial hemorrhage | **I62** |
| Cerebral aneurysm | **I67.1** |
| Transient ischaemic attack (TIA) | **G45** |
| **Deep vein thrombosis (DVT)** | **I80** |
| **Pulmonary embolism (PE)** | **I26** |
| **Other venous embolism and thrombosis** | **I82** |
| **Arrhythmias** |  |
| Atrial fibrillation and flutter | **I48** |
| Other cardiac arrhythmias | **I49** |

## MATERIALS AND METHODS

### Study summary

**Table 2** provides the summary of the study information.

**Table 2.**
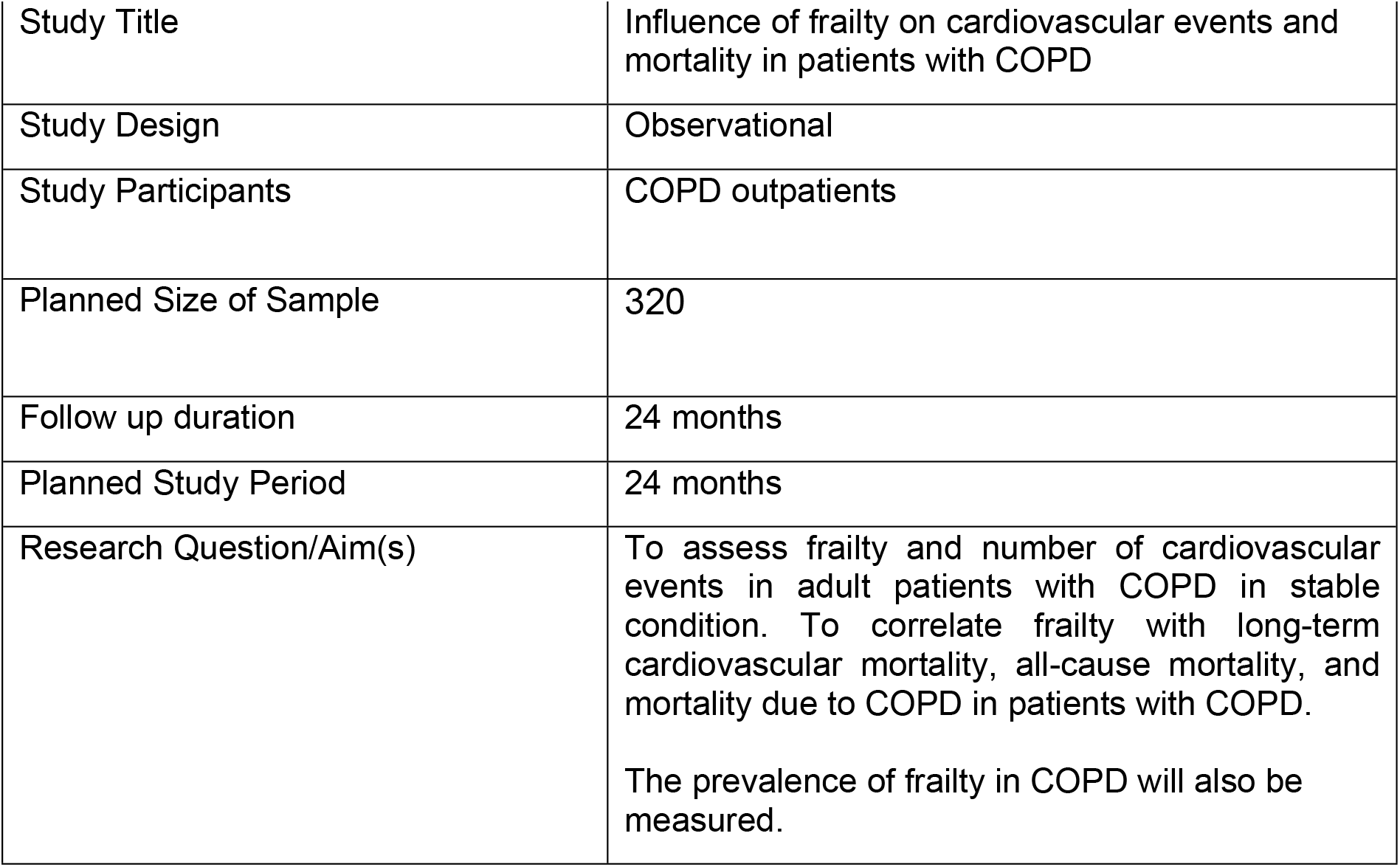
Study summary.

### Study design and setting

#### Study design

Multicentre European observational study. The study follows the STROBE statement for reporting of cohort studies.

#### Study setting

Two university hospitals and one local health care service in Italy, and university hospital site in the UK that provide care and follow up/scheduled visits for outpatients diagnosed with COPD will participate. The central study team subsequently provided the ethical approval, protocol, central organisation and long-term delivery of the project.

### Study population

#### Patient screening

Patients will be screened for inclusion criteria by the local team in the outpatient setting at each hospital site. The diagnosis of COPD will be according to the GOLD guidelines (www.goldcopd.org) and confirmed by a post-bronchodilator FEV_1_/FVC < 0.70.

Principal Investigators (PIs) at each site will have local responsibility for data quality and entry. Data collection will be carried out using the case report form (CRF) presented in the Supporting Information file S1. Where personal information is collected at each site, it will be kept secure, and maintained. This will involve the creation of a number code for each patient (Patient Identification, PID) that will guarantee data anonymisation.

#### Inclusion criteria

Patients with age ≥18 years, with diagnosis of COPD of at least 1 year, smoking history of ≥ 10 pack/years, in stable condition, without acute exacerbation in the last 30 days and/or all-cause hospitalisation in the last 3 months, able to provide their written informed consent, will be included.

#### Exclusion criteria

Terminally ill patients, patients with concurrent diagnosis of asthma, concurrent diagnosis of interstitial lung disease, presence of restrictive pattern on spirometry, no history of smoking habit, not able to provide their written informed consent, will be excluded.

### Power and sample size

Assuming the proportion of those that are not frail (40%), mildly frail (60%) and moderately to severely frail (75%) will exhibit at least one cardiovascular event at 1 year. In order to detect these two comparisons (not frail, versus mildly frail & not frail versus moderately frail), using a chi-squared test with type-1 error=0.025 and 80% power. We will need a minimum of 320 patients, approximately 120:120:80 for CFS 1-3, 4-5, 6-8 respectively.

### Variables

A SPIRIT schedule of enrollment, interventions, and assessments is reported in **Figure 1**.

**Figure 1.**
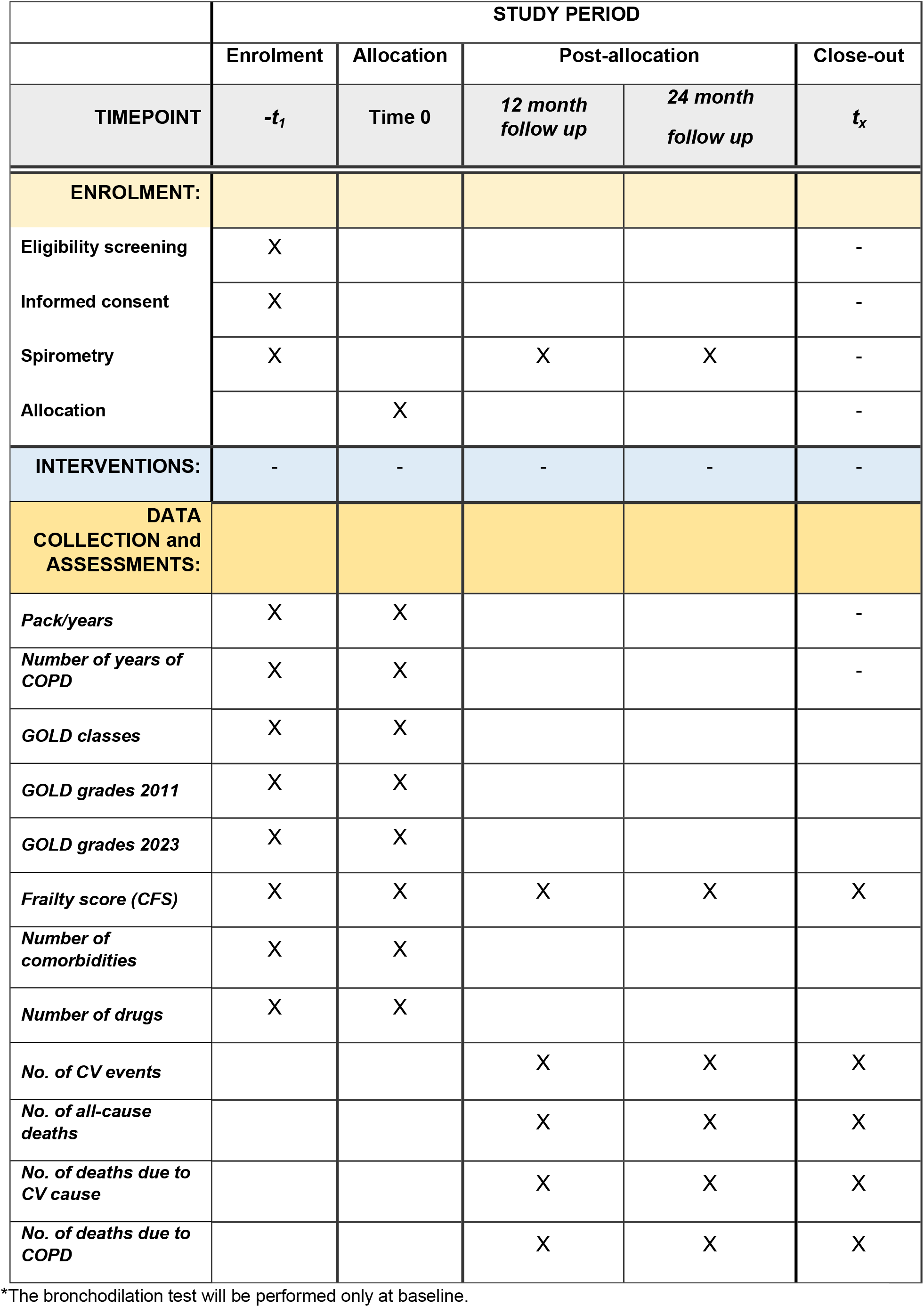
SPIRIT schedule of enrolment, interventions, and assessments.

### Baseline data

➢ Age, sex, occupation (current or previous), body mass index (BMI, Kg/M^2^), active smoking expressed as pack/years (= number of cigarettes per day multiplied by the number of years the person has smoked/20), years of COPD, GOLD classes (1, 2, 3, 4) and GOLD grades based on GOLD 2011 (A, B, C, D) and GOLD 2023 reports (A, B, E) (www.goldcopd.org), frailty score using the Clinical Frailty Scale (**Figure 2**), list and number of comorbidities, list and number of drugs taken.

### Cardiovascular events

These will be defined as the following. Any ICD-10 code reported in **Table 1** or the following event recorded in the hospital notes at 1 and 2 years will be considered as a cardiovascular event.

**Figure 2.**
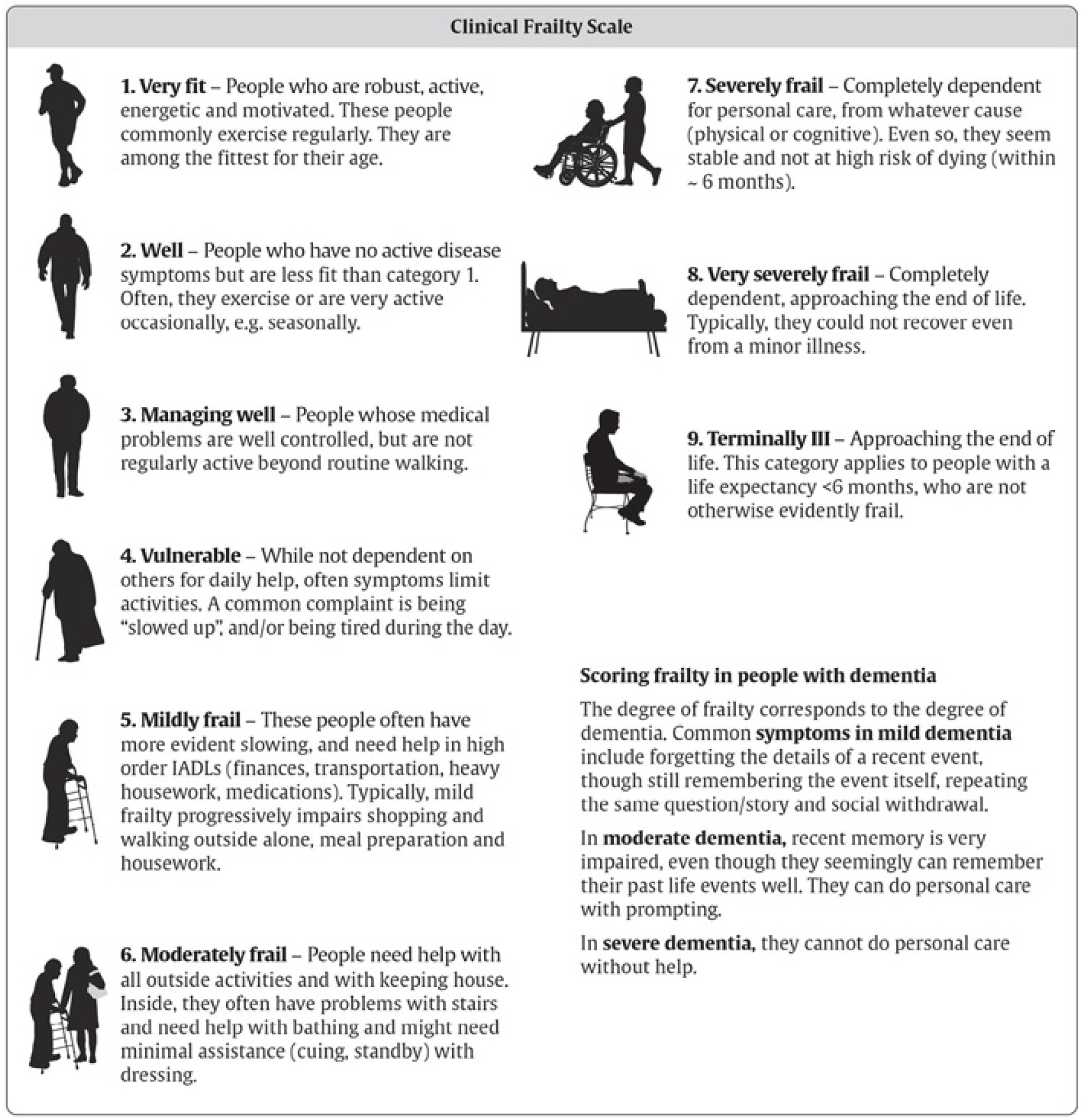
Clinical Frailty Scale.

### Follow up data (12 and 24 months)

➢ Number of cardiovascular events.
➢ Number of deaths due to cardiovascular cause.
➢ Number of all-cause deaths.
➢ Number of deaths due to COPD.
➢ Frailty score using the Clinical Frailty Scale (**Figure 2**).
➢ Follow up spirometry.

### Frailty assessment

We will use the Clinical Frailty Score. This has been validated for use to assess frailty in older patients. [17] The score ranks from 1 to 9 with a score of ≥4 being classed as mildly frail, and 4-5 and moderately frailty, and 6 to 9 severely frail to terminally ill. Patients scored CFS 9 (terminally ill) will be excluded.

### Safety considerations

Safety reporting/adverse events will be reported by Principal Investigators. However, as this study routinely collected clinical data, there are minimal safety issues expected.

### Primary outcome

➢ Number of cardiovascular events (including hospital admissions due to cardiovascular cause) at 12 and 24 months in COPD patients living with frailty *vs* non-frail COPD patients.

### Secondary outcomes

➢ Long-term cardiovascular mortality at 12 and 24 months in COPD patients living with frailty *vs* non-frail COPD patients.
➢ All-cause long-term mortality at 12 and 24 months in COPD patients living with frailty *vs* non-frail COPD patients.
➢ Mortality due to COPD at 12 and 24 months in COPD patients living with frailty *vs* non-frail COPD patients.
➢ Prevalence of frailty in COPD.

### Data management plan, data protection and patient confidentiality

All investigators and trial site staff will comply with the requirements of the Good Clinical Practice (GCP) and Guidelines for Data Processing in the European Union (General Data Protection Regulation-GDPR 679/2016) with regards to the collection, storage, processing and disclosure of personal information.

Where personal information is collected, it will be kept secure, and maintained. This will involve:

- The creation of coded, depersonalised data where the participant’s identifying information is replaced by an unrelated sequence of characters.
- No patient identifiable information will be uploaded or stored on the secure database. Collaborators will anonymise patients by recording patient clinical numbers alongside database numbers in a separate secure spreadsheet to aid the collection of data locally.
- Limiting access to the minimum number of individuals necessary for quality control, audit, and analysis.
- Confidentiality of data will be preserved when the data are transmitted to co-investigators as patients will be assigned a study ID number.
- Data collected from sites will be via the electronic database, using freely available software such as Microsoft XL. The research team are experienced in data management. This database with be held within University of Modena (Italy) and stored for a minimum of 7 years.
- Data custodian and Chief Investigator is Dr Alessia Verduri.
- Only researchers who are directly involved in the data analysis will have full access to the study dataset. Participating sites may request access to the full dataset from the Chief Investigator. The research team will have full access to the study dataset in order to ensure that the overall results are not disclosed by an individual trial site prior to the main publication.

### Statistical analysis

We will aim to recruit a minimum of 320 patients: across a minimum of 3 sites.

This protocol and analysis plan will be drafted by a medical statistician fully blinded to the outcome data.

Data will be analysed for correlation between frailty and clinical outcomes. The primary analysis will be the number of cardiovascular events (time to event) at 12 and 24 months, adjusting for sex, age (18-64, 65-80, >80) and frailty at the time of recruitment, and other clinically relevant non-respiratory comorbidities. The number of events will be analysed using a zero-inflated Poisson regression to compare the rates between the three frailty groups. The rate with 95%CI will be presented alongside the p-value. Overdispersal will be assessed and if found to be present robust standard errors will be fitted.

The secondary analyses will assess the time to all-cause mortality, cardiovascular mortality, and mortality due to COPD that will be fitted to compare those classified as frail *vs* not frail visually using a Kaplan Meier plot. Data will be analysed using a multivariable Cox baseline proportional hazards regression, adjusting for covariates. The time to mortality will be calculated from the date of study entry to the end of follow up. The analyses will be presented as Odds Ratio (adjusted OR) with associated 95% confidence intervals and p-values. Subgroups analyses (age, sex, GOLD classes and grades, comorbidities) will be conducted to assess the grade of frailty. Stata version 18 (or later) will be used.

### Validation Data

Validation will be performed by local teams on 25% of data fields for 10% of cases. The validated fields will include key demographic and outcome data.

### Patient and Public Involvement (PPI) Group

It will be possible to develop a PPI group of patients who have participated to the study. Towards the end of the study, the coordinator centre will invite a group of patients back to the hospital to discuss their personal opinion about the meaning of the research study.

### Ethics

The study was approved and registered by the University of Modena and Reggio Emilia Ethics Committee Area Vasta Nord Emilia-Romagna (Modena, Italy); approval number 448/2023/OSS/AOUMO – SIRER ID 6390, 14^th^ November 2023. The study was opened at each participating site. All participating units must obtain approval from their local Ethics Committee. It is the responsibility of the local study team to ensure that the ethical approvals are in place prior to commencing data collection. The principal investigators at each site will ensure that collaborators act in accordance with local clinical governance and guidelines. The study has been externally peer reviewed by impartial professionals with relevant expertise (respiratory physicians, geriatricians, statisticians).

In the UK, this study was deemed to be a service evaluation, individually approved for use in the UK hospital site.

The study has been registered within www.clinicaltrials.gov: NCT05922202.

All the participants will provide written informed consent (Supporting Information file S1).

### Status and timeline of the study

Recruitment of participants began on the 19^th^ December 2023 at the Hospital Policlinico in Modena (Italy) and will continue for two years (20^th^ December 2025).

## DISCUSSION

This protocol describes a study designed to investigate the influence of frailty on the risk of cardiovascular morbidity and mortality in patients with COPD. The study will also assess whether the frailty status can have an impact on all-cause mortality and death due to COPD. This study will offer key real world data components to describe the complexity of COPD that is frequently associated with cardiovascular disease or other concomitant chronic diseases sharing common risk factors and playing a role in COPD prognosis. Additionally, patients with COPD have high risk of frailty that can increase their risk of mortality. [7-12]

Relevant data collected includes comorbidities and GOLD grades based on the GOLD recommendations 2011 and 2023 (www.goldcopd.org) to characterise patient’s severity related to risk of exacerbation, hospitalisation, and death. The primary research questions this study will answer are whether the frailty status may be associated with the number of cardiovascular events and long-term cardiovascular mortality in a large, complete population with COPD. Answers to the questions of this longitudinal cohort study will inform chest physicians of the role of frailty in patients with COPD. More broadly, the clinical data collected in outpatient setting, including frailty, can have significant value to predict patient’s prognosis. This is a current topic in the field of respiratory research.

One limitation of the study design is the lack of a control group of individuals matched on age, sex, smoking habit, and frailty status, but without COPD. This would be useful to distinguish the potential role of COPD in the risk of development of cardiovascular disease [18].

All data will be reported as a whole cohort. Unit-level data for comparison will be fed back to collaborators to support local service improvement. This project will be submitted for presentation at a national or international Respiratory and Geriatric conference. Manuscript(s) will be prepared following close of the project. Manuscript(s) will also be prepared following interim data analysis at 12 months if numbers of patients recruited to the study at that time reach the planned sample size.

## Data Availability

No datasets were generated or analysed during the current study. All relevant data from this study will be made available upon study completion.

## Authors’ contributions

**Conceptualisation:** Alessia Verduri

**Investigation:** Alessia Verduri, Enrico Clini, Jonathan Hewitt

**Study Design:** Alessia Verduri, Enrico Clini, Ben Carter, Jonathan Hewitt

**Statistical considerations**: Ben Carter

**Project administration:** Alessia Verduri, Enrico Clini

**Supervision:** Alessia Verduri, Enrico Clini, Ben Carter, Jonathan Hewitt

**Writing – original draft:** Alessia Verduri

**Writing – review & editing:** Alessia Verduri, Enrico Clini, Ben Carter, Jonathan Hewitt.

## Acknowledgements

Not applicable.

## Supporting Information

The SPIRIT checklist, informed consent form, and case report form (CRF) are presented in the Supporting Information file S1.

